# Assessing the Incidence of Postoperative Diabetes in Gastric Cancer Patients: A Comparative Study of Roux-en-Y Gastrectomy and Other Surgical Reconstruction Techniques

**DOI:** 10.1101/2024.01.13.24301276

**Authors:** Tatsuki Onishi

**Author notes:** Corresponding author: Onishi Tatsuki.

## Abstract

**Study objective:** Sleeve gastrectomy is effective in morbid obesity, and it improves glucose homeostasis. In gastric cancer patients with type 2 diabetes mellitus, gastrectomy, including total gastrectomy (TG), is beneficial for glycaemic control. However, the effects of gastrectomy and different reconstructive techniques on the incidence of postoperative diabetes in gastric cancer patients are unclear. This study investigated the development of new-onset diabetes in these patients, focusing on different reconstruction methods.

**Design:** A comparative study

**Setting:** Electrical medical records

**Patients:** This study included 715 patients without diabetes who underwent TG at Tokyo Metropolitan Bokutoh Hospital between August 2005 and March 2019.

**Interventions:** Patients underwent reconstruction by Roux-en-Y (RY) or other surgical techniques (OT), with diabetes onset determined by HbA1c levels or medical records.

**Measurements:** Analyses included two-sample t-tests, chi-squared tests, and the Kaplan-Meier method with log-rank tests to compare the onset curves between the two groups.

**Main Results:** Stratified data analysis compared the RY and OT reconstruction methods. Log-rank test results (P=0.0217) indicated a statistically significant difference in the incidence of new-onset diabetes between RY and OT groups in gastric cancer patients.

**Conclusion:** This first-of-its-kind study provides insight into how different methods of gastric reconstruction affect postoperative diabetes. The results suggest significant differences in new-onset diabetes mellitus after surgery based on the reconstruction method. This research highlights the need for careful surgical planning to consider potential postoperative diabetes, particularly in patients with a family history of diabetes mellitus. Future studies should investigate the role of gut microbiota and other reconstructive techniques, such as laparoscopic jejunal interposition, in developing postoperative diabetes.

## 1. Introduction

Gastrectomy, particularly sleeve gastrectomy (SG), has been shown to be an effective surgical option for morbid obesity due to its low complication rates and significant weight loss results [1–5]. SG results in alteration of the appetite through the regulation of gut hormones, resulting in decreased hunger and increased satiety [6]. SG also improves glucose homeostasis through resulting changes in gut hormone levels [7]. Specifically, laparoscopic SG results in significant improvement in glucose metabolism in morbidly obese patients and has been found to stop the development of diabetes at a high rate [8]. SG has been shown to improve blood glucose independently of weight loss by restoring hepatic insulin sensitivity [9]. However, the effects of gastrectomy on non-obese patients with type 2 diabetes are less clear, with some studies suggesting that gastrectomy may improve diabetic status [10].

In patients with gastric cancer diagnosed with type 2 diabetes mellitus (T2DM), gastrectomy has been found to have a positive impact on their glycemic control. Improvements in glycemic control, or even diabetes remission, have been reported after gastrectomy [10–15]. The extent of the gastrectomy, duration of diabetes, and method of reconstruction have been identified as important factors influencing the improvements in glycemic control [10–14]. Although the mechanisms underlying these effects are not fully understood, oncometabolic surgeries, including gastrectomy, have been suggested as a potential treatment for T2DM in patients with gastric cancer [16].

Studies have shown that total gastrectomy (TG) is associated with improved glucose metabolism in patients with gastric cancer, resulting in a lower rate of newly diagnosed diabetes after surgery [22]. However, the effects of gastrectomy on glucose metabolism in diabetic and non-diabetic patients have been inconsistent, with some studies reporting significant reductions in fasting blood glucose levels after gastrectomy [23]. Furthermore, SG has been associated with significant reductions in HbA1c levels in non-diabetic patients, suggesting its possible role in the prevention of type 2 diabetes [24].

In terms of reconstruction after partial gastrectomy in patients with gastric cancer, both Roux-en-Y and Billroth II reconstructions have been considered acceptable options [17]. Roux-en-Y reconstruction is often preferred for gastric cancer patients, given that this procedure can lead to decreased reflux gastritis and esophagitis, decreased probability of cancer recurrence, and decreased incidence of surgical complications [18]. Roux-en-Y reconstruction has also been found to be as effective as other methods with respect to nutritional status and postoperative outcomes [19]. In comparison to Billroth II reconstruction, Roux-en-Y has been shown to have similar postoperative complications and better long-term outcomes [20]. Furthermore, Roux-en-Y reconstruction without cutting has been the preferred method in cases of gastritis, bile reflux, and gastric residuals [21].

Various studies have examined the impact of different reconstructive procedures on postoperative complications in gastric cancer patients. It has been found that long-limb Roux-en-Y bypass reconstruction could lead to improved glycemic control [25], and it has been observed that pre-existing diabetes is associated with postoperative complications [10, 26]. Several studies further support the benefits of Roux-en-Y reconstruction, with some indicating it to be more effective than Billroth II reconstruction [27, 28]. Additionally, significant improvements in diabetes control have been associated with Roux-en-Y reconstruction [25, 28].

Given these, the aim of this research was to investigate the incidence of new-onset diabetes in patients with gastric cancer after surgery and how this incidence varies with different types of surgical reconstruction, namely, the Roux-en-Y procedure and other alternative reconstruction techniques. While studies have investigated how surgical treatment for gastric cancer affects existing diabetes [25, 28], none have investigated the development of new-onset diabetes in patients without diabetes; to the best of our knowledge, this is the very first study to do so. Findings from this study could contribute valuable insights into the postoperative outcomes associated with different gastric reconstruction techniques. Such insights are vital for guiding clinical decisions and optimizing patient care, particularly in the context of mitigating the risk of developing diabetes after gastric surgery. Moreover, findings from this study are expected to have significant implications for both clinical practice and future research in the field of gastric surgery and diabetes prevention.

## 2. Materials and Methods

### 2.1 Study participants

A total of 715 patients who underwent TG as the definitive procedure and as a standby procedure at the Tokyo Metropolitan Bokutoh Hospital between August 2005 and March 2019 and were not diagnosed with diabetes at the time of surgery were included in the study (Table 1). The study was approved by the institution’s ethics committee (30-110) and conducted in accordance with the Declaration of Helsinki.

**Table 1.**
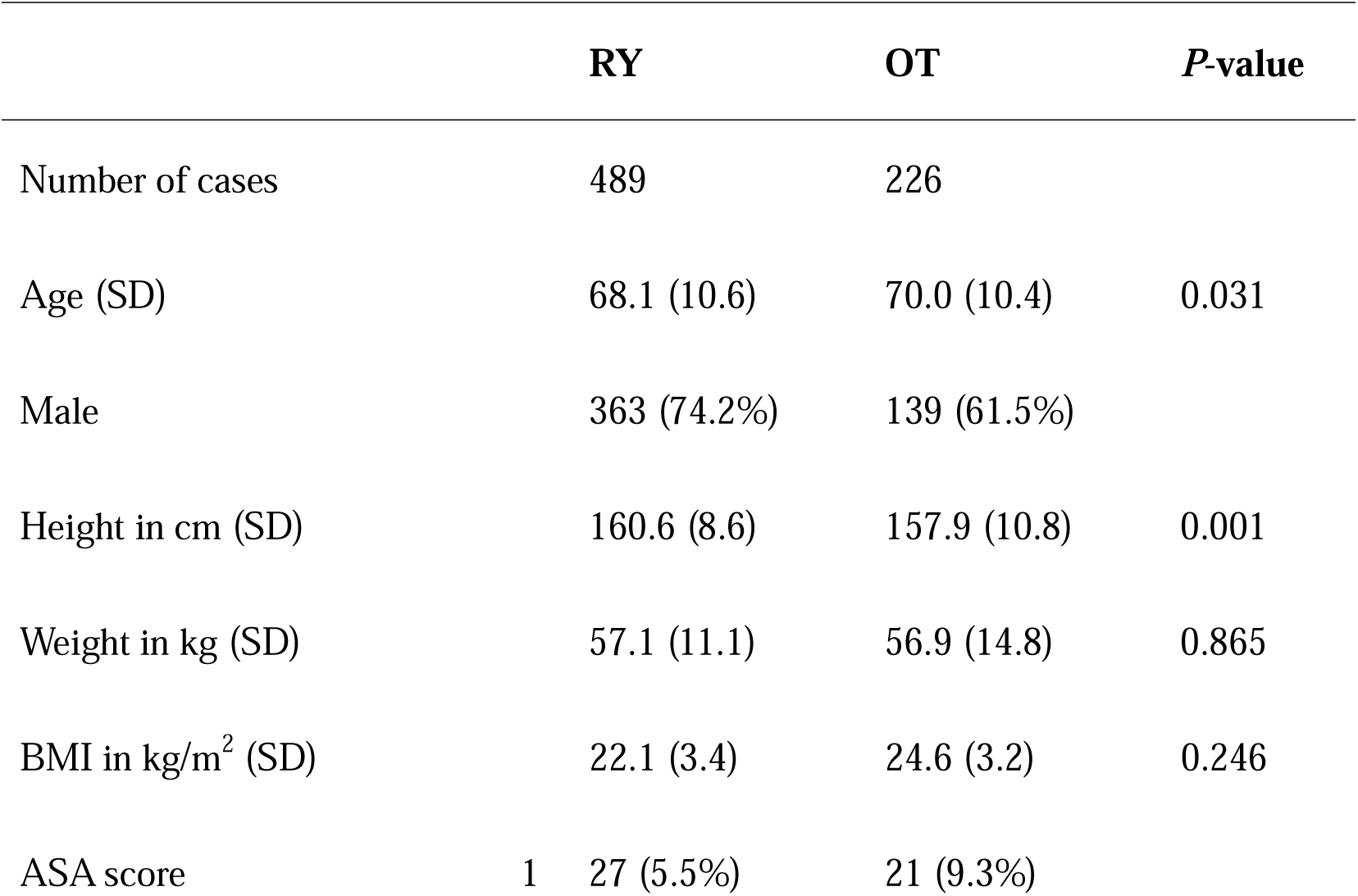
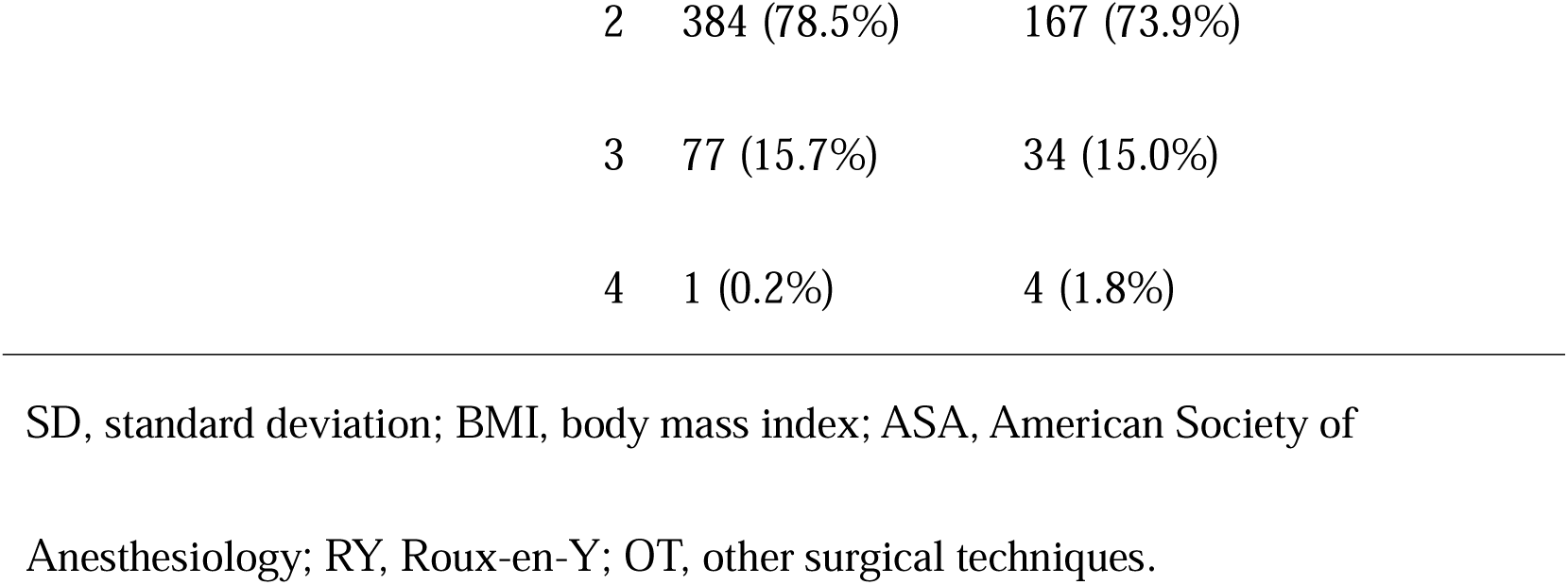
Demographics of the study population.

### 2.2 Methods

Whether the patients would undergo reconstruction through Roux-en-Y (RY) or other surgical techniques (OT) was chosen based on previous studies, and the patients were grouped accordingly. The definite onset of diabetes in the patients was considered based on the previous electronic medical records or when their HbA1c value was greater than 6.5 based on laboratory testing. The competing outcome was death. After a meticulous data curation process using Python 3.10 that corrects for missing values, manages outliers, and ensures appropriate data types, we obtained a data set that was optimized for analysis and free of common data inconsistencies.

Furthermore, basic statistical measures such as the mean, median, and standard deviation were computed using Python 3.10. Two sample t-test and Chi-squared test for variables in characteristics were used to assess the difference in demographics of the two groups using Python 3.10. In addition, the Kaplan–Meier method was employed to estimate the onset function from the time-event data present and was augmented with log-rank tests to help compare the onset curves between the RY and OT groups using Python 3.10.

## 3. Results

The characteristics of the patients included in the study at the time of the surgery are shown in Table 1.

In the present study, following rigorous data pre-processing, a Kaplan–Meier onset curve was constructed to estimate the function delineating the interval between the total gastrectomy and the subsequent emergence of new-onset diabetes postoperatively (Fig. 1). This nonparametric approach provides a survival function, articulated as a step function, which quantifies the probability of patients remaining free from diabetes up to a specific time point following surgery.

**Figure 1:**
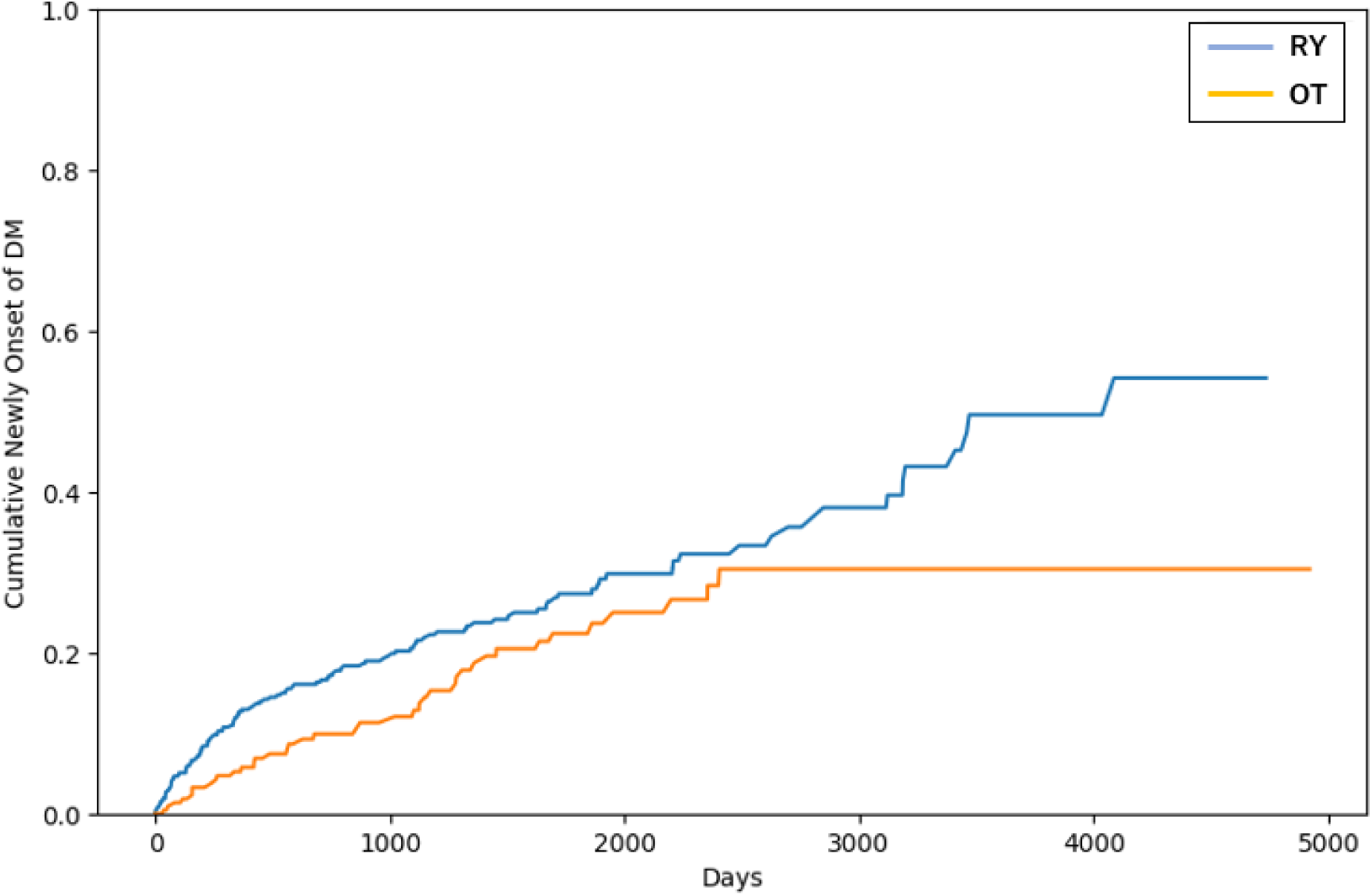
Kaplan-Mayer curve of newly onset of DM in RY and OT group RY, Roux-en-Y; OT, other surgical techniques.

The analytical framework was further refined by stratifying the data to draw comparisons between the two types of reconstruction methods applied, the RY and OT. This stratification was imperative for a granular comparison of the incidence rates of postoperative diabetes associated with these distinct reconstructive procedures. Kaplan–Meier curves were employed for each stratified group, offering a visual representation of the temporal pattern of diabetes onset post-surgery corresponding to each surgical technique. The rate of diabetes onset was inferred from the slope of these curves, with a steeper decline indicating a higher incidence within the respective group.

To statistically ascertain the significance of the disparities observed in the incidence curves between the groups, a log-rank test was conducted. The resultant *P-*value from this log-rank test was found to be 0.0217, which denotes a statistically significant difference in the incidence of new-onset diabetes post-surgery between those who underwent RY and those subjected to OT in patients with gastric cancer. These findings indicate a difference in the incidence of postoperative diabetes based on the type of gastric reconstruction method employed.

## 4. Discussion

This study is the first to provide insights into how different methods of gastric reconstruction might affect the risk of developing postoperative diabetes. This analysis is important for understanding the temporal dynamics of diabetes development after gastric surgery and has significant implications for surgical planning and patient management to prevent postoperative diabetes. A thorough approach to data pre-processing and the use of robust statistical methods will ensure the reliability and validity of these findings in the wider context of gastric surgery and diabetes research.

Although the results of this study show that RY is associated with a higher incidence of new-onset postoperative diabetes compared with OT, these data could be improved by comparison with a population that has not undergone total gastrectomy as a control. Nevertheless, the finding that new-onset diabetes mellitus after surgery differs according to the method of gastric cancer reconstruction should be an important consideration in surgical selection, especially in cases with a strong family history of diabetes mellitus.

Surgery for gastric cancer, particularly gastrectomy, has been shown to significantly alter the gut microbiota, leading to dysbiosis characterized by changes in bacterial content and gene function [31, 32]. This dysbiosis is associated with intestinal inflammation, overgrowth of small intestinal bacteria and an increased risk of colorectal cancer [32]. Specific changes in the gut microbiota after surgery include increased species richness, decreased butyrate-producing bacteria, and enrichment of certain symbiotic bacteria [33]. The abundance of specific gut bacterial genera has been found to correlate with the population of peripheral immune cells [33–35].

This study did not include an assessment of other determinants that could potentially influence the development of diabetes mellitus, including lifestyle choices and genetic predisposition. It is plausible that there may be a difference in the intrinsic characteristics of diabetes mellitus in patients who present with diabetic symptoms prior to undergoing surgery for gastric cancer, as opposed to those in whom the onset of gastric malignancy precedes the development of diabetes mellitus. Such considerations were beyond the scope of analysis within the parameters of the current study.

Similarly, laparoscopic jejunal interposition (LJIP) reconstruction, a surgical technique used to treat gastric cancer in which a pouch is created in the jejunum and used to reconstruct the upper gastrointestinal tract, may be appropriate for patients with impaired glucose tolerance [36]. Studies have shown that LJIP reconstruction leads to better postoperative outcomes, including improved quality of life and nutritional status, compared with other reconstruction methods, such as double tract reconstruction [36–39]. The procedure has also been successfully performed laparoscopically with promising results in terms of operative time, blood loss and postoperative recovery [40, 41].

In this study, LJIP reconstruction was not performed at the institution, and it was not possible to study the gut microbiota. With access to a suitable dataset, we would like to investigate the association between gut microbiota and the development of new-onset diabetes after surgery further based on different reconstructive methods, including LJIP reconstruction.

## Acknowledgements

Tatsuyoshi Ikenoue at Shiga University, Data Science and AI Innovation Research Promotion Center for proof reading, Yoshika Onishi at Wellbeing Keiei LLC for proof reading

## Data availability statement

Data is available upon request to the corresponding author. Codes have been uploaded here https://github.com/bougtoir/gastric_dm

## Author contributions

O.T.: conceptualization, writing - original draft, data curation, visualization, and final approval of the manuscript.

